# Impact of Treatment Adherence and Inhalation Technique on asthma outcomes of Pediatric Patients: A Longitudinal Study

**DOI:** 10.1101/2023.11.30.23299186

**Authors:** Catalina Lizano-Barrantes, Olatz Garin, Karina Mayoral, Alexandra L. Dima, Angels Pont, M. Araceli Caballero-Rabasco, Manuel Praena-Crespo, Laura Valdesoiro-Navarrete, María Teresa Guerra, Alberto Bercedo-Sanz, Montse Ferrer, the ARCA Group

## Abstract

**Introduction:** We aimed to evaluate the longitudinal relationships, both at between- and within-person levels, that adherence to inhaled corticosteroids-based maintenance treatment and inhalation technique present with symptom control, exacerbations, and health-related quality of life (HRQoL) in children and adolescents with asthma.

**Methods:** Participants (6-14 years old) from the ARCA (Asthma Research in Children and Adolescents) cohort - a prospective, multicenter, observational study (NCT04480242) - were followed for a period from 6 months to 5 years, via computer-assisted telephone interviews and a smartphone application. The Medication Intake Survey–Asthma (MIS-A) was administered to assess the implementation stage of adherence; and the Inhalation Technique Questionnaire (InTeQ) to assess the five key steps when using an inhaler. Symptoms control was measured with the Asthma Control Questionnaire (ACQ), and HRQL with the EQ-5D and the PROMIS-Pediatric Asthma Impact Scale (PROMIS-PAIS). Multilevel longitudinal mixed models were constructed separately with symptom control, exacerbation occurrence, EQ-5D, and PROMIS-PAIS as dependent variables.

**Results:** Of 360 participants enrolled, 303 (1203 interviews) were included in the symptom control and exacerbation analyses, 265 (732) in the EQ-5D, and 215 (619) in the PROMIS-PAIS. Around 60% of participants were male and most underwent maintenance treatment with inhaled corticosteroids plus long-acting β-agonists in a fixed dose (68–74%). Within-person variability was 83.6% for asthma control, 98.6% for exacerbations, 36.4% for EQ-5D and 49.1% for PROMIS-PAIS. At within-person level, patients with higher adherence had better symptom control (p=0.002) and HRQoL over time (p=0.016). Patients with better inhalation technique reported worse HRQoL simultaneously (p=0.012), but better HRQoL in future assessments (p=0.012). Frequency of reliever use was associated with symptom control (p<0.001), exacerbation occurrence (p<0.001), and HRQoL (p=0.042); and boys were more likely to present better symptom control and HRQoL than girls.

**Conclusion:** Our results confirm longitudinal associations at within-person level of the two indicators of quality use of inhalers: for adherence to maintenance treatment with symptom control and HRQoL, and for inhalation technique with HRQoL. Although treatment adherence showed to be excellent, a third part of participants reported a suboptimal inhalation technique, highlighting the need of actions for improving asthma management of pediatric population.

## 1. Introduction

Asthma is the most common non-communicable disease in school-aged children [1,2] and is a major public health problem worldwide [1,3,4]. In 2019, an estimated 12,900 deaths occurred and 5.1 million disability-adjusted life years were lost due to childhood asthma [5]. According to the latest global report, only 44.1% of children and 55.4% of adolescents with asthma achieved a well-controlled disease [6].

Childhood asthma is a heterogeneous and fluctuating disease, with symptoms that vary in time and intensity [7,8]. Therefore, management is mainly based on a continuous personalized cycle of assessment of asthma control (symptom control and risk factors for future exacerbations), any comorbidities that could contribute to symptom burden and poor Health-Related Quality of Life (HRQoL), and treatment [7]. Daily inhaled corticosteroids (ICS) is the currently recommended pharmacologic maintenance therapy in individuals of all ages [7,9]. Research has shown that adherence to ICS and inhalation techniques are dynamic and complex [10–12], with studies indicating generally low adherence to maintenance medication (20-70%) [13,14] and suboptimal inhalation technique (8– 22%) [15] in children and/or adolescents.

Most evidence from systematic reviews suggests whether it is children [16–20], adolescents [17–22], or adults [17–19,21–23], higher levels of adherence [17,21,23] and better inhalation technique [18,19,22], analyzed separately, are associated with better outcomes (symptom control, exacerbations and/or HRQoL), although an inverse or null association has also been found [17–19,21–23]. Furthermore, impaired HRQoL has also been linked with asthma-associated factors, such as severity [16], disease control, and exacerbations [20,21] in children, adolescents, and young adults.

These systematic reviews [17–23] show that more than 160 studies have been conducted involving patients with asthma that evaluated the relationships between adherence, inhalation technique, asthma control, asthma exacerbation, and/or HRQoL. However, only 22 of these studies included longitudinal analyses, with 9 focusing exclusively on children and/or adolescents [13,24–31] and 13 encompassing adults as well [32–44]. Notably, none of them considered the temporal stages of adherence (initiation, implementation, and discontinuation) described in 2012 [45]. Although the systematic reviews were published between 2015 and 2022, none of the studies including longitudinal analyses were conducted after 2013. Therefore, they are probably not reflecting the Food and Drug Administration (FDA) requirement changes, contraindicating the use of long-acting beta-agonists (LABA) without concurrent ICS [46].

The Global Asthma Initiative Guideline (GINA) [7] has continued to incorporate changes due to the collection of new evidence related to the efficacy and safety of ICS, LABA, and short-acting beta-agonists (SABA). More recent longitudinal studies [10,47–54] have presented further evidence on the long-term role of ICS adherence in asthma. However, none were specifically on pediatric population, only one included HRQoL [51], few specified the adherence stage considered [47,48,53,54], and only one included medication adherence alongside with inhalation technique [51]. These two last concepts are closely related, with the poor inhalation technique even being considered an unintentional form of adherence [55], but they are usually identified as independent concepts [56]. Two of the aforementioned systematic reviews [17,18] have highlighted the scarcity of studies evaluating the impact of adherence and inhalation technique, assessed together, on asthma outcomes, despite the association that has been observed between them [57,58].

A deeper insight into how adherence and inhaler technique evolve over time, and affect clinical outcomes and HRQoL in children, could foster a ’quality use of medications’ strategy [59], aligning with current guidelines. Therefore, we aimed to evaluate the longitudinal relationships, both at between- and within-person levels, that adherence to ICS (alone or in combination with LABA) and inhalation technique present with symptom control, exacerbations, and HRQoL in children and adolescents with asthma.

## 2. Materials and methods

### Study design and participants

The Asthma Research in Children and Adolescents (ARCA) is a longitudinal, prospective, multicenter, observational study (NCT04480242), designed to provide evidence about the evolution of young patients with persistent asthma through regular follow-up.

Patients were consecutively recruited in 5 outpatient pediatric pulmonology hospital units and 9 primary care pediatric centers in Spain, from January 2018 to March 2023, thus followed for a period from 6 months to 5 years. Inclusion criteria were: aged 6–14, with a clinical diagnosis of asthma, undergoing treatment with inhaled corticosteroids (alone or combined with LABA) for more than 6 months in the previous year, no concomitant respiratory diseases, and with access to a smartphone (their own or their parents’). Written informed consent was requested from parents or legally authorized representatives of all participants, and additionally, oral consent was obtained from children.

Participants were followed *via* the ARCA smartphone app [60] monthly, and *via* computer-assisted telephone interviews performed by trained interviewers at enrolment, every 6 months (regular CATIs), and after each exacerbation (post-exacerbation CATIs). The ARCA app is available in 3 age versions: proxy response for 6–7 years old children, and self-response for 8–11 and for ≥ 12 years old participants. Through the app, participants reported any new exacerbations and completed HRQoL instruments. Two versions of the CATIs were administered, one for parents or guardians of children under 8 years old (proxy response) and one for participants aged 8 and older (self-response). CATIs collected information on asthma symptom control, exacerbations, asthma treatments (maintenance and reliever), adherence to maintenance medication, inhalation technique, reliever use, and exacerbation occurrence, for the period immediately before the interview. Demographic and clinical information was collected from medical records at enrolment.

For this analysis, we selected participants who had valid registries of at least two CATIs during a period with an ICS-based treatment prescribed for regular use (maintenance).

The ESPACOMP Medication Adherence Reporting Guideline (EMERGE) was followed [84].

### Study variables

Medication information was collected at every CATI administered to participants. Maintenance treatment was grouped into two categories: ICS in fixed-dose combination with LABA (ICS plus LABA) and single ICS inhaler. The frequency of reliever medication use was measured with the following question: *How often have you usually taken your “reliever medication” (brand name) in the past 4 weeks: every day; almost every day; once or twice every week; or less than once a week?* This variable was grouped into: Almost never (participants with no SABA prescribed and those reporting *used less than once a week*); and Usually (participants reporting the first 3 response options).

Medication adherence was measured with the Medication Intake Survey–Asthma (MIS-A) [61], a validated instrument for telephone interviews, which assesses the implementation stage of adherence separately for each maintenance inhaler based on self-reported prescription start date, daily dosage recommendations, and questions on maintenance use over increasing periods. Percentages of used versus prescribed medication are calculated first for each question and subsequently as composite scores. We used 1-month composite scores based on inhalations used the day before (Q1); days on which no inhalations were taken in the past 7 days (Q2); days on which all prescribed inhalations were used in the past 7 days (Q3); and days on which all prescribed inhalations were used in the past 28 days month (Q4). The MIS-A was administered at enrolment and every 6 months in the regular CATIs, and also in the post-exacerbation CATIs. When patients used more than one inhaler containing ICS, we computed scores for each inhaler and averaged across them. The MIS-A has been validated [61] using self-response in adult patients and teenagers, and a proxy version for caregivers of children.

The inhalation technique was measured with the Inhaler Technique Questionnaire (InTeQ) [62,63], an instrument that assesses the frequency of performing five key steps when using the inhaler in the previous six months with a five-level Likert scale (from “Always” to “Never”). The InTeQ was administered in the CATIs at enrolment and yearly. A global score was calculated as a sum of the InTeQ items answered "Always", among the four which demonstrated unidimensionality in children and adolescents [63], and was categorized into: 4–3 (Good inhaler technique), 2 (Fair), and 1–0 (Poor). The InTeQ has been validated for telephone interviews [63] using self-response in children aged 8 and older and proxy response for parents or guardians of children under 8 years old. As the InTeQ was only administered yearly, missing values were replaced by data from the previous interview.

Symptom control was measured with the Asthma Control Questionnaire (ACQ) – symptoms only [64], which was administered in the regular and post-exacerbation CATIs. It assesses the presence and intensity of night-time waking, symptoms on waking, activity limitation, shortness of breath, and wheezing during the previous week on a 7-level Likert scale from 0 (no impairment) to 6 (maximum impairment). The overall score, calculated as the mean item responses, ranges from 0 to 6. Cut-off points of 1.5 and 0.75 were established to define not well- and well-controlled asthma, respectively [65]. The ACQ has been validated [66] using self-administration in adolescents and interviewer-administration in children.

Asthma exacerbations were identified in the regular CATIs administered every 6 months or by reporting them through the app, which prompted an alert to the research team that was followed by a post-exacerbation CATI to confirm its occurrence. In both cases, exacerbations were defined through three questions constructed applying the definitions by the American Thoracic Society and the European Respiratory Society [67]: *Did you visit or phone your family doctor or outpatient emergency department because your asthma got worse? Did you call an ambulance or go to the hospital because of your asthma? Did you take steroid tablets or syrup (such as Prednisolone or Deltacortril) for at least 2 days because of your asthma?* If the participant answers “yes” to at least one of the three questions, an asthma exacerbation is confirmed.

Health-related quality of life (HRQoL) was measured using 2 complementary instruments: the EuroQol generic questionnaire (EQ-5D) [68–70] and the disease-specific questionnaire (Patient-Reported Outcomes Measurement Information System-Pediatric Asthma Impact Scale) PROMIS-PAIS [71], which were administered through the ARCA app. The EQ-5D was administered at enrolment and every 6 months. It consists of 5 dimensions: “mobility”, “looking after myself”, “doing usual activities”, “having pain/discomfort” and “feeling worried/sad/unhappy, with a time frame of “today”. According to age, we used the EQ-5D-Y-3L proxy-version (6–7 years), the self-administered EQ-5D-Y-3L (8–11 years), and the self-administered EQ-5D-5L (≥ 12 years). A single preference-based utility index was calculated ranging from 1 (the best health state) to negative values (health states valued by society as worse than death), where 0 is equal to death. Preference value sets applied to generate this utility index were those obtained from Spanish adults for the EQ-5D-5L [72] and those obtained from Spanish adults thinking as a hypothetical 10-year-old child for the EQ-5D-Y [73,74]. The short form 8a version of the PROMIS-PAIS (v2.0) was administered at 4 months from enrolment and every 6 months thereafter. Its items ask about the past seven days in a 5-level Likert response scale (1–5) with the options: never, almost never, sometimes, often, and almost always. It is available for self-response in ages 8–17 and for proxy response in children starting at age 5. The total raw score is calculated by adding the values of the response to each question, ranging from 8 to 40 (lower score indicates better HRQoL) [75].

### Analytical strategy

To specifically examine the impact of the implementation stage of adherence to an ICS-based maintenance treatment (i.e., the degree to which patients follow their prescribed doses during treatment), we censored from the dataset reports under certain conditions: no prescribed daily ICS at all, ICS prescribed on an as-needed basis, or prescribed other asthma maintenance treatment (such as tiotropium). Descriptive analyses were performed of patient characteristics, treatment, and outcomes by calculating percentages, or means and standard deviations. Differences between the patients included and not included in the analysis of each outcome (asthma symptom control, exacerbation, EQ-5D, and PROMIS-PAIS) were assessed with chi-square or t-test, according to the type of variable.

Continuous time-varying predictors (adherence and inhalation technique) were decomposed into three variables to distinguish the between-person effects and the simultaneous and sequential within-person effects. Average adherence was calculated as the mean score for each patient across all reports (one score per patient) and used for examining whether differences in adherence between patients predict outcomes. Current fluctuation was computed as the difference between a patient’s average adherence and their score in a given report (multiple scores per patient) to examine whether changes in adherence within patients are associated with concomitant changes in outcome (i.e. measured in the same report). Prior fluctuation was computed as lagged variable, i.e. the difference between a patient’s average and the score in their previous report, usually 6 months earlier (multiple scores per patient) to examine whether changes in adherence predict outcomes measured in the subsequent report.

To assess longitudinal relationships of adherence to ICS-based maintenance treatment and inhalation technique with outcomes, we followed established procedures for hierarchical longitudinal modeling [76]. Four multilevel longitudinal mixed models were constructed separately for asthma symptom control, exacerbation occurrence, EQ-5D, and PROMIS-PAIS (as dependent variables). In all cases, models were constructed to assess the role of the two time-varying variables, adherence and inhalation technique (which are the main explanatory variables), including them together with the type of ICS-based maintenance treatment, and sociodemographic variables that can be potential confounders (Model A); then adding other factors that are part of the implicit standard for asthma management [7,77]: use of reliever, asthma symptom control, and the occurrence of exacerbations, except in models where they were the dependent variable (Model B). Time was modeled as years since the first interview per patient (random and fixed) and interactions between the independent variables and time were tested. Further than p values of each coefficient or odds ratio (OR) provided by the models, ANOVA were applied to test the significance corresponding to each independent variable. Sensitivity analyses were performed with 1-week adherence scores. R (version 4.2.2), and RStudio (2022.07.2 Build 576) were used to construct all the models, except for the exacerbation occurrence, which was constructed with SAS 9.4.

## 3. Results

Out of the 360 participants enrolled from January 2018 to March 2023 (Fig. 1), we excluded from the analysis: 10 who did not respond any CATI, 42 with only one valid CATI, and 5 without ICS-based maintenance treatment. Then, 303 participants (1,203 CATIs) were included in the asthma symptom control and exacerbation analysis, 265 participants (732 CATIs) in the EQ-5D, and 215 (619 CATIs) in the PROMIS-PAIS. In the analyses of symptom control and exacerbation occurrence, patients provided 2-9 reports (median 4, IQR [2-5]), with a mean (SD) follow-up of 492 (419) days (range 116-1759 days). For the EQ-5D and PROMIS-PAIS analyses, reports per patient ranged from 1-8, with medians of 2 (IQR [1-3]) and 3 (IQR [2-4]), respectively.

**Figure 1.**
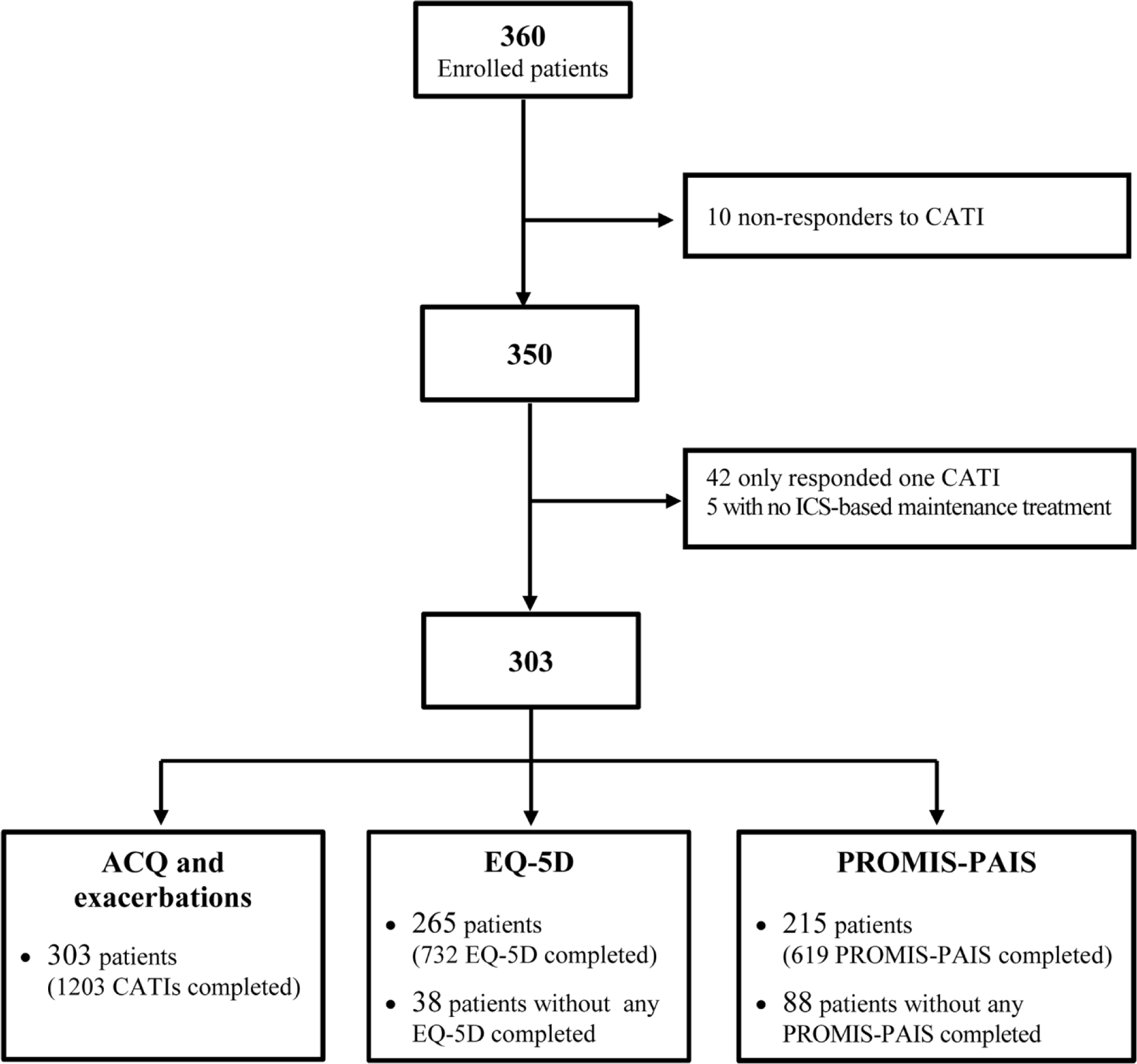
Flowchart for the selection of patients meeting analysis criteria. CATI: computer-assisted telephone interview; ICS: Inhaled Corticosteroids; ACQ: Asthma Control Questionnaire; PROMIS-PAIS: PROMIS-Pediatric Asthma Impact Scale.

The characteristics of participants are presented in Table 1. The majority were male (approximately 60%), aged between 8 and 11 years (48-51%), undergoing maintenance treatment with inhaled corticosteroids combined with LABA in a fixed dose (73%), and reported almost never using relievers (approximately 61%). The mean 1-month adherence scores were around 88%, around 45% of participants reported good inhalation technique, and more than 64% had well-controlled symptoms. Experiencing exacerbations was reported by around 38% of the participants. The HRQoL score measured with the EQ-5D was 0.93 (1 -best health state- to negative values -worse than death), and measured with the PROMIS-PAIS it was 13.4 (8 -best health state- to 40 -worst).

**Table 1.** Demographic and clinical characteristics of participants.

|  | All<br>(n=350) | ACQ and<br>Exacerbations<br>(n=303) | EQ-5D<br>(n=265) | PROMIS-PAIS<br>(n=215) |
| --- | --- | --- | --- | --- |
| **Sex, n (%)** |  |  |  |  |
| **Male** | 216 (61.7%) | 184 (60.7%) | 160 (60.4%) | 130 (60.5%) |
| **Female** | 134 (38.3%) | 119 (39.3%) | 105 (39.6%) | 85 (39.5%) |
| *p*-value |  | 0.334 | 0.364 | 0.544 |
| **Age, n (%)** |  |  |  |  |
| **6 – 7 years** | 86 (24.6%) | 73 (24.1%) | 65 (24.5%) | 52 (24.2%) |
| **8 – 11 years** | 166 (47.4%) | 147 (48.5%) | 131 (49.4%) | 110 (51.2%) |
| **≥12 years** | 98 (28.0%) | 83 (27.4%) | 69 (26.0%) | 53 (24.7%) |
| *p*-value |  | 0.586 | 0.297 | 0.142 |
| **Maintenance treatment, n (%)** |  |  |  |  |
| **ICS** | 101 (28.9%) | 81 (26.7%) | 72 (27.2%) | 56 (26.0%) |
| **ICS plus LABA** | 241 (68.9%) | 222 (73.3%) | 193 (72.8%) | 159 (74.0%) |
| **Other treatment** | 7 (2.0%) | 0 (0.0%) | 0 (0.0%) | 0 (0.0%) |
| **No treatment** | 1 (0.3%) | 0 (0.0%) | 0 (0.0%) | 0 (0.0%) |
| *p*-value |  | 0.000 | 0.000 | 0.001 |
| **Reliever use, n (%)** |  |  |  |  |
| **Not prescribed** | 22 (6.3%) | 19 (6.3%) | 18 (6.8%) | 14 (6.5%) |
| **Less than once a week** | 194 (55.4%) | 168 (55.4%) | 147 (55.5%) | 114 (53.0%) |
| **Once or twice a week** | 90 (25.7%) | 80 (26.4%) | 70 (26.4%) | 60 (27.9%) |
| **Almost every day** | 29 (8.3%) | 25 (8.3%) | 23 (8.7%) | 21 (9.8%) |
| **Every day** | 14 (4.0%) | 10 (3.3%) | 6 (2.3%) | 5 (2.3%) |
| *p*-value |  | 0.648 | 0.094 | 0.175 |
| *Missing* | 1 | 1 | 1 | 1 |
| **% Last month adherence, mean (SD)** | 86.7 (23.3) | 87.8 (21.3) | 88.0 (21.0) | 88.4 (20.8) |
| *p*-value |  | 0.029 | 0.056 | 0.084 |
| *Missing* | 23 | 15 | 11 | 10 |
| **% Last week adherence, mean (SD)** | 85.0 (27.2) | 85.8 (25.9) | 86.7 (25.1) | 86.4 (25.5) |
| *p*-value |  | 0.125 | 0.032 | 0.198 |
| **Inhalation technique, n (%)** |  |  |  |  |
| **Poor (0 – 1 Always)** | 106 (31.7%) | 95 (32.5%) | 83 (32.0%) | 65 (31.0%) |
| **Fair (2 Always)** | 81 (24.3%) | 65 (22.3%) | 56 (21.6%) | 48 (22.9%) |
| **Good (3 – 4 Always)** | 147 (44.0%) | 132 (45.2%) | 120 (46.3%) | 97 (46.2%) |
| *p*-value |  | 0.082 | 0.094 | 0.556 |
| *Missing* | 16 | 11 | 6 | 5 |
| **ACQ, mean (SD)** | 0.8 (1.1) | 0.8 (1.1) | 0.8 (1.0) | 0.8 (1.0) |
| *p*-value |  | 0.781 | 0.185 | 0.327 |
| **Not well controlled (> 1.5)** | 71 (20.9%) | 63 (21.3%) | 51 (19.5%) | 40 (18.9%) |
| **Intermediate (0.75 - 1.5)** | 48 (14.2%) | 43 (14.5%) | 40 (15.3%) | 35 (16.5%) |
| **Well controlled (< 0.75)** | 220 (64.9%) | 190 (64.2%) | 171 (65.3%) | 137 (64.6%) |
| *p*-value |  | 0.767 | 0.328 | 0.185 |
| *Missing* | 11 | 7 | 3 | 3 |
| **Exacerbation, n (%)** |  |  |  |  |
| **No** | 210 (61.9%) | 184 (62.2%) | 161 (61.5%) | 134 (63.2%) |
| **Yes** | 129 (38.1%) | 112 (37.8%) | 101 (38.5%) | 78 (36.8%) |
| *p*-value |  | 0.830 | 0.728 | 0.537 |
| *Missing* | 11 | 7 | 3 | 3 |
| **EQ-5D** |  |  | 0.93 (0.11) |  |
| **PROMIS** |  |  |  | 13.0 (5.7) |

p-values assessing differences between the patients included and those not included in each subsample correspond to chi-square or t-test, according to the type of variable.

Table 2 shows results for the longitudinal associations that maintenance treatment adherence and inhalation technique present with asthma symptom control (left column) and exacerbations (right column). The proportion of between-person variation was 16.4% for asthma control, and 1.4% for exacerbations. Model A with asthma symptom control shows that, at within-person level, patients reporting higher adherence to maintenance medication also reported better control in the next interview (prior fluctuation; p=0.002). On the contrary, both models A and B show that girls (p=0.006 and p=0.012) were more likely to report worse control of asthma symptoms. Furthermore, Model B shows that patients who reported using reliever medication ≥1-2 times per week (p<0.001) and having an exacerbation (p<0.001) were also more likely to present uncontrolled asthma symptoms. Age, the inhalation technique, and the type of maintenance treatment (ICS alone or in combination with LABA) did not present statistically significant association to asthma symptom control.

**Table 2.** Multilevel models of asthma symptom control (linear) and exacerbation occurrence (logistic)

|  |  | Asthma symptom control |  | Exacerbation occurrence |  |
| --- | --- | --- | --- | --- | --- |
|  |  | Model A | Model B | Model A | Model B |
|  |  | b (SE) |  | OR (SE) |  |
| **Intercept** |  | 0.281 (0.253) § | 0.069 (0.203) § | 0.45 (0.60) | 0.81 (0.69) |
| **Time (years)** |  | 0.000 (0.003) | 0.000 (0.002) | 0.97 (0.01) *** § | 0.97 (0.01) *** § |
| **ADHERENCE** |  |  |  |  |  |
| Average adherence |  | 0.001 (0.003) | 0.001 (0.002) | 1.00 (0.01) | 0.99 (0.01) |
| Current fluctuation of adherence |  | -0.002 (0.002) | -0.002 (0.001) | 0.99 (0.00) | 0.99 (0.00) |
| Prior fluctuation of adherence |  | -0.005 (0.002) ** § | -0.002 (0.001) § | 0.99 (0.00) | 0.99 (0.01) |
| **INHALATION TECHNIQUE** |  |  |  |  |  |
| Average IT |  | 0.044 (0.040) | 0.016 (0.032) | 1.21 (0.10) | 1.16 (0.10) |
| Current fluctuation of IT |  | 0.002 (0.036) | -0.009 (0.033) | 1.10 (0.12) | 1.06 (0.12) |
| Prior fluctuation of IT |  | -0.032 (0.040) | -0.046 (0.036) | 1.14 (0.13) | 1.11 (0.14) |
| **Treatment** |  |  |  |  |  |
| ICS plus LABA |  | Ref. | Ref. | Ref. | Ref. |
| ICS |  | 0.120 (0.091) | 0.073 (0.074) | 1.06 (0.23) | 0.97 (0.24) |
| **Sex** |  |  |  |  |  |
| Male |  | Ref. § | Ref. § | Ref. | Ref. |
| Female |  | 0.233 (0.085) ** | 0.168 (0.067) * | 1.06 (0.20) | 0.89 (0.21) |
| **Age** |  |  |  |  |  |
| < 8 years |  | Ref. | Ref. | Ref. § | Ref. § |
| 8 – 11 |  | -0.097 (0.104) | -0.055 (0.083) | 0.44 (0.24) *** | 0.42 (0.25) *** |
| ≥12 |  | -0.053 (0.119) | -0.009 (0.094) | 0.38 (0.28) *** | 0.34 (0.30) *** |
| **Reliever use** |  |  |  |  |  |
| Almost never |  |  | Ref. |  | Ref. § |
| Usually |  |  | 0.840 (0.063)<br>*** § |  | 3.27 (0.23) *** |
| **Exacerbation** |  |  |  |  |  |
| No |  |  | Ref. |  |  |
| Yes |  |  | 0.354 (0.067)<br>*** § |  |  |
| **Asthma symptom control** |  |  |  |  |  |
| Not well controlled |  |  |  |  | Ref. § |
| Intermediate |  |  |  |  | 0.42 (0.38) * |
| Well controlled |  |  |  |  | 0.47 (0.29) ** |
| ICC (linear); VPC (logistic) |  | 0.2568 | 0.1641 | 0.0129 | 0.0142 |
| logLikelihood |  | -1085.3 | -978.8 |  |  |
| AIC |  | 2200.6 | 1991.5 |  |  |
| BIC |  | 2271.0 | 2071.4 |  |  |
| -2 Res Log Pseudo-Likelihood |  |  |  | 3726.66 | 3838.27 |
| Generalized Chi-Square |  |  |  | 678.05 | 665.65 |
| Generalized Chi-Square / DF |  |  |  | 0.87 | 0.86 |

The p-values corresponding to each coefficient or OR provided by the models were marked with asterisks: \*(p<0.05); \*\*(p<0.01); \*\*\*(p<0.001). ANOVA p-values for each independent variable were marked with § (p<0.05).
ICS: Inhaled Corticosteroids; ICS plus LABA: Inhaled Corticosteroids plus Long-Acting Beta-Agonists; ICC: Intraclass Correlation Coefficient; VPC: Variance Partition Coefficients; AIC: Akaike Information Criterion; BIC: Bayesian Information Criterion.

Exacerbations Models A and B show less risk of occurrence in children aged 8 years or older (p≤0.001 in both models) and participants reporting better asthma symptom control (p= 0.023 and p= 0.008). Conversely, risk of exacerbation occurrence is higher in participants reporting using reliever medication ≥1-2 times per week (p<0.001). Neither average adherence and inhalation technique, nor their prior or simultaneous fluctuations were associated with exacerbation occurrence.

The proportion of between-person variation was 63.6% and 50.9%% for HRQoL (Table 3), EQ-5D and PROMIS-PAIS, respectively. The EQ-5D models reveal that, when participants reported better inhalation technique, they reported worse HRQoL simultaneously (current fluctuation; p=0.012 and p= 0.012), but better HRQoL in the next interview (prior fluctuation; p= 0.005 and p= 0.012). Furthermore, worse HRQoL was more likely in girls (p=0.037 and p= 0.036). Age, adherence, type of treatment, the use of reliever medication, and the occurrence of exacerbations were not statistically significantly associated with EQ-5D.

**Table 3.** Multilevel models of Health-Related Quality of Life measured with the EQ-5D and PROMIS-PAIS (linear)

|  |  | EQ-5D |  | PROMIS-PAIS |  |
| --- | --- | --- | --- | --- | --- |
|  |  | Model A | Model B | Model A | Model B |
|  |  | b (SE) |  | b (SE) |  |
| **Intercept** |  | 0.986 (0.044) *** § | 0.988 (0.046) *** § | 11.264 (2.384) *** § | 11.988 (2.397) *** § |
| **Time (years)** |  | 0.000 (0.001) | 0.000 (0.001) | -0.034 (0.028) | 0.038 (0.140) |
| **ADHERENCE** |  |  |  |  |  |
| Average adherence |  | 0.0004 (0.000) | 0.0004 (0.000) | 0.027 (0.024) | 0.031 (0.023) |
| Current fluctuation of adherence |  | 0.0002 (0.000) | 0.0002 (0.000) | 0.007 (0.012) | 0.010 (0.012) |
| Prior fluctuation of adherence |  | 0.0004 (0.000) | 0.0002 (0.000) | -0.023 (0.012) | -0.009 (0.013) |
| **INHALATION TECHNIQUE** |  |  |  |  |  |
| Average IT |  | 0.002 (0.007) | 0.003 (0.006) | -0.624 (0.379) | -0.658 (0.352) |
| Current fluctuation of IT |  | -0.015 (0.006) * § | -0.015 (0.006) * § | -0.316 (0.328) | -0.244 (0.324) |
| Prior fluctuation of IT |  | 0.021 (0.007) ** § | 0.019 (0.007) * § | -0.231 (0.364) | -0.094 (0.359) |
| **Treatment** |  |  |  |  |  |
| ICS plus LABA |  | Ref. | Ref. | Ref. | Ref. |
| ICS |  | -0.012 (0.016) | -0.011 (0.016) | -0.074 (0.902) | -0.041 (0.847) |
| **Sex** |  |  |  |  |  |
| Male |  | Ref. § | Ref. § | Ref. § | Ref. § |
| Female |  | -0.029 (0.014) * | -0.029 (0.014) * | 2.538 (0.824) ** | 2.457 (0.764) ** |
| **Age** |  |  |  |  |  |
| < 8 years |  | Ref. | Ref. | Ref. § | Ref. § |
| 8 - 11 |  | 0.010 (0.017) | 0.006 (0.017) | -0.395 (0.979) | 0.073 (0.918) |
| ≥ 12 |  | 0.005 (0.020) | 0.001 (0.020) | 2.383 (1.169) * | 2.709 (1.093) * |
| **Asthma symptom control** |  |  |  |  |  |
| Not well controlled |  |  | Ref. § |  | Ref. § |
| Intermediate |  |  | -0.018 (0.019) |  | 0.305 (1.053) |
| Well controlled |  |  | 0.016 (0.014) |  | -2.403 (0.825) ** |
| **Reliever use** |  |  |  |  |  |
| Almost never |  |  | Ref. |  | Ref. |
| Usually |  |  | -0.012 (0.012) |  | 1.319 (0.646) * |
| **Exacerbation** |  |  |  |  |  |
| No |  |  | Ref. |  | Ref. |
| Yes |  |  | -0.014 (0.011) |  | 0.082 (0.616) |
| **Interaction time \* Adherence last month** |  |  |  |  |  |
| Average Adherence |  |  |  |  | -0.001 (0.002) |
| Current fluctuation of adherence |  |  |  |  | 0.001 (0.001) |
| Prior fluctuation of adherences |  |  |  |  | -0.003 (0.001) * § |
| ICC |  | 0.6161 | 0.6362 | 0.5716 | 0.5090 |
| logLikelihood |  | 312.7 | 304.6 | -1386.4 | -1400.3 |
| AIC |  | -591.4 | -567.3 | 2838.2 | 2848.6 |
| BIC |  | -522.5 | -482.3 | 2907.7 | 2946.3 |

The p-values corresponding to each coefficient provided by the models were marked with asterisks: \*(p<0.05); \*\*(p<0.01); \*\*\*(p<0.001).
ANOVA p-values for each independent variable were marked with § (p<0.05).
ICS: Inhaled Corticosteroids; ICS plus LABA: Inhaled Corticosteroids plus Long-Acting Beta-Agonists; ICC: Intraclass Correlation Coefficient; VPC: Variance Partition Coefficients; AIC: Akaike Information Criterion; BIC: Bayesian Information Criterion.

In PROMIS-PAIS models the interaction between time and adherence reveals an increase in HRQoL over time, correlating with higher levels of patient-reported adherence in subsequent interviews (prior fluctuation; p=0.016). Furthermore, better asthma symptom control was also associated with better HRQoL (p= 0.004). Conversely, worse HRQoL was more likely for adolescents compared to children under 12 years of age (p= 0.043 and p=0.014), girls (p= 0.001 and p=0.001), and the use of reliever medication ≥1-2 times a week (p=0.042). The type of maintenance treatment regimen, the inhalation technique, and exacerbation did not present a statistically significant association with PROMIS-PAIS.

Sensitivity analysis with 1-week adherence scores showed similar results (supplementary material).

## 4. Discussion

This study provides evidence regarding the longitudinal relationships that maintenance treatment adherence and inhaler technique present with asthma symptom control, exacerbations occurrence, and HRQoL in pediatric asthma patients. We gathered comprehensive patient-reported data using a combination of the ARCA app and CATIs. We found that better asthma symptom control over time (future assessments) was more likely in patients with higher adherence to treatment, while boys and those participants reporting almost never using reliever medication or no exacerbations generally had better symptom control. In the same direction, lower risk of exacerbations was found in older children, those reporting well controlled symptoms and almost never using reliever medication. Better HRQoL over time was observed in patients who reported better adherence and inhalation technique. Additionally, boys and participants with better symptom control generally had better HRQoL.

### Adherence to ICS-based maintenance treatment

The level of adherence to maintenance treatment in ARCA participants is high, on average: they reported having administered 88% of prescribed dose during the previous month, which is above the range of 20-70% identified by a systematic review [13,14] in children and/or adolescents.

Consistent with our hypothesis, we found that higher adherence was associated with better asthma symptom control in future assessments, despite the inconsistent results reported both by systematic reviews [16,22], which mainly included cross-sectional studies, and by more recent longitudinal studies [48,49,53,54]. Consistently with our finding, a longitudinal study in French and English adults and children with asthma [48] showed that patients maintaining high ICS adherence over time have better asthma control. In the same line, a study of the large Nivel Primary Care Database in the Netherlands shows association between poor ICS adherence and uncontrolled asthma [53]. Conversely, a UK study [54] using the Optimum Patient Care Research Database (OPCRD) found that patients might adjust their ICS based on current needs without this necessarily impacting later in hospitalizations, emergency visits, outpatient visits, or the need for oral corticosteroids or antibiotics. Additionally, a longitudinal study in patients from 27 countries with ICS plus LABA maintenance treatment pointed out that most patients only use medication when they are not well [49]. Overall, these findings lead us to incorporate nuances into our hypothesis: the association between adherence and asthma control might be driven by increased adherence as a reactive response to uncontrolled symptoms, which could eventually lead to increased symptom control over time.

The association found between increased treatment adherence and increased HRQoL over time is also consistent with our hypotheses, as it could reflect an individual’s overall investment in maintaining their health and well-being through effective asthma management practices. This association was particularly identified with the asthma-specific questionnaire PROMIS-PAIS, likely due to its focused content, potentially more responsive to asthma symptoms [78]. While the specific association of adherence with HRQoL has been less frequently examined, our results are consistent with findings of a systematic review in adolescents [20] and a multicenter, observational, prospective study in Greek adults with variable asthma severity [79]. Additionally, a longitudinal study in Dutch adolescents [30] indicated that higher HRQoL at baseline predicted increased medication adherence at follow-up, although good medication adherence did not predict an increase in HRQoL over time. These results line up with our enhanced hypothesis, distinguishing patients with regular adherence, who actively integrate treatment into their daily routines, recognizing its importance, from those with “reactive adherence”, who strictly follow treatments only when they feel that their asthma is out of control.

Nevertheless, our findings indicate a lack of association between adherence and exacerbation occurrence. Although they were consistent with observations from the abovementioned longitudinal studies in France, UK and the Netherlands [48,53,54], they contrast with results of a meta-analysis [80] indicating that higher treatment adherence reduces exacerbation risk, while discontinuation increases it. It is important to highlight that response bias cannot be discarded in our study since interviews were performed immediately after experiencing an exacerbation, possibly making patients feel accountable. In fact, the concept of accountability for their treatment adherence, which refers to the influence on patient behavior of the expectation of social interactions with healthcare providers, has been poorly explored in previous research [81].

### Inhalation technique

In our study, 32% of participants reported poor inhalation technique, which is above the proportion of suboptimal inhalation technique reported by the studies of children and/or adolescents with asthma (8– 22%) identified in a systematic review [15].

Given the recognized importance of both inhalation technique and adherence in impacting actual drug exposure [7,77], we hypothesized finding a similar association when both factors were analyzed together. Our results focusing on within-person fluctuations of inhalation technique revealed that when participants temporarily improved their technique, their HRQoL decreased during that same period, but improved afterwards. This is also consistent with our hypothesis distinguishing between regular and reactive behaviors, suggesting similar patterns for inhalation technique and adherence, where a proactive approach to asthma management, even if initially challenging, ultimately contributes to enhanced HRQoL. These fluctuations are likely due to factors changing within patients with asthma over time, rather than stable differences between patients, as highlighted in the longitudinal study involving French and English adults and children [48].

Three systematic reviews [18,19,22] supported that better inhalation technique, analyzed without considering adherence, is consistently associated with exacerbations [19,22] and HRQoL [18,19,22], but there are less consistent results with asthma symptoms control [18,19,22]. However, evidence on the relationship between inhalation technique and HRQoL remains limited. For instance, one of the reviews [18] included one single prospective longitudinal clinical study with a small sample size. Another one [19] referenced only two intervention-focused studies to enhance inhalation technique. The third review [22] exclusively referenced a cross-sectional study assessing HRQoL, which found no significant outcome differences between patients based on inhalation technique. This state of the art highlights the need for further comprehensive research to fully understand the impact of inhalation technique on various asthma-related outcomes. The lack of a statistically significant association between inhalation technique and the other outcomes of our study deserves further research.

### Frequency of reliever use

Our findings about the association of frequent use of reliever medication with uncontrolled asthma symptoms and exacerbation occurrence align with those from the Nivel Primary Care Database from Netherlands [53], which also observed them. Two studies conducted across European countries [82] and Canada [83] also reported the association between the use of SABA and exacerbations occurrence. Furthermore, our study revealed an association between frequent reliever use and worse HRQoL, a relationship that has been less explored. A cross-sectional analysis of the study in France and UK measuring the impact of asthma [84] showed statistically significant differences on HRQoL according the frequency of reliever medication use; among women, those using reliever medication almost or every day presented the biggest deviation from reference norms.

Our findings suggest that the frequent use of reliever medication, again potentially reflecting a reactive approach to asthma management, negatively impacts HRQoL. This observation ties in with our earlier hypothesis regarding adherence and inhalation technique, where proactive self-management practices are contrasted with reactive behaviors. Such patterns underline the complex dynamics of asthma self-management and emphasize the need for future research to conduct a more in-depth exploration of the within-person fluctuations in reliever use and its impact on HRQoL.

### Asthma symptom control

The positive long-term association between asthma symptom control and HRQoL found in our study was consistent with a longitudinal study in dyads of asthmatic children and their parents in USA [85] showing that poorly controlled asthma status was associated with poor HRQoL. Additionally, a systematic review on adolescents [21] identified poor disease control, exacerbations, and asthma severity as main factors associated with impaired HRQoL. Apparently in contrast, the longitudinal Dutch study in adolescents [30] found that higher HRQoL at baseline did not predict changes in asthma control over time. On the other hand, the lower risk of exacerbations among patients with better asthma symptom control observed in our study aligns with the Asthma Care logic process Model [77] and the GINA guideline [7], which position asthma control as directly related to exacerbations.

### Sociodemographic factors

Our research identified gender differences in asthma outcomes, with girls experiencing worse asthma symptom control and HRQoL compared to boys. This finding is supported by literature reviews [21,86,87] that also link an association of HRQoL and asthma control impairment with female gender. Additionally, we observed that individuals aged 12 years and older showed a decreased HRQoL. These associations could be attributed to hormonal changes impacting airway inflammation, potential variances in immune responses, and the distinctive psychosocial challenges faced by females and adolescents as previously pointed out [86–88]. These factors might collectively contribute to worsened asthma symptoms and treatment outcomes, subsequently affecting HRQoL. Nevertheless, the Dutch study in adolescents [30] observed an increase in adolescents’ HRQoL over time, attributing this to the possibility that they may perceive their illness as less of a concern. Furthermore, we found that a lower risk of exacerbations was associated with a higher age, which could be related to fewer virus-induced exacerbations, since they are more common in younger children [89].

### Limitations

Interpreting our findings requires taking into account various limitations. First, we didńt consider the interplay of other important factors such as comorbidities (rhinitis, obesity, and anxiety, among others) and environmental triggers. Second, our results don’t preclude the potential benefits of a deliberate effort to improve overall adherence and inhalation techniques due to the participation in a study, which could potentially impact their relationship with outcomes, and the outcomes themselves. Third, the InTeQ’s reliance on a long recall period (previous 6 months) introduces a potential recall bias. And fourth, our analysis did not differentiate among LABAs in the ICS fixed-dose combination treatments as established by guidelines [7]. Unfortunately, our sample size disbalance between users (69 reports for ICS-formoterol, 712 ICS-salmeterol, and 124 ICS-vilanterol) prevented carrying out stratified analysis to explore any differences. Finally, measurement of adherence and inhalation technique are based on patient or proxy reporting. Therefore, future research studying these complex relationships could benefit from pharmacy claims, performance tests, and smart inhalers.

## 5. Conclusions

To our knowledge, this is the first longitudinal study specifically conducted in pediatric patients to assess both HRQoL and clinical outcomes (asthma symptom control and exacerbation occurrence), allowing for the evaluation of their longitudinal relationships with two of the main indicators of quality use of inhalers (i.e., adherence and inhalation technique). Methodologically, the hierarchical mixed model approach adopted has the advantage of describing how each person changes over time (within-person), and how these changes differ across people (between-person). In addition, conceptually, the Timelines-Events-Objectives-Sources (TEOS) framework [93] has been applied to operationalize adherence.

Our findings highlight the multifaceted nature of asthma in children and adolescents, getting closer to a comprehensive understanding of the dynamic process of asthma treatment and outcomes over time. It is remarkable how, although treatment adherence showed to be excellent, a third part of participants reported a suboptimal inhalation technique, supporting the need of actions for improvement in the asthma management of pediatric population. We found longitudinal associations at within-person level of the two indicators of quality use of inhalers: for adherence to ICS-based maintenance treatment with symptom control and HRQoL, as well as for inhalation technique with HRQoL. This reinforces the importance of further examining changes over time, alongside changes across people. Notably, frequency of reliever use was associated with symptom control, exacerbation occurrence, and HRQoL; pointing out the need of examining within-person changes in reliever use, further than the usually assessed between-person differences. Finally, due to the differences observed between boys and girls, it is especially important to apply a gender perspective in clinical practice and future studies on children and adolescents with asthma.

## Author Contributions

CLB contributed to conceptualization, investigation, methodology, project administration, visualization, writing—original draft, review and editing. MF contributed to conceptualization, funding acquisition, methodology, project administration, supervision, visualization, writing–review and editing. OG and KM contributed to conceptualization, methodology, visualization, writing–review and editing. ALD contributed to methodology, writing–review, and editing. AP contributed to data curation, formal analysis, and writing–review and editing. MACR, MPC, LVN, MTG, and AB contributed to the investigation and writing–review and editing. All the co-authors critically revised the manuscript and approved the submitted version.

## Data Availability

All data produced in the present study are available upon reasonable request to the authors.

## Acknowledgments

Authors would like to thank the participants of the study. They also thank Áurea Martín for her support in English editing and proofreading. ARCA Group: Montse Ferrer, Karina Mayoral, Catalina Lizano, Olatz Garin, Yolanda Pardo, Àngels Pont (Hospital del Mar Research Institute); Maria Araceli Caballero (Hospital del Mar); Manuel Praena (Centro de Salud la Candelaria); Laura Valdesoiro (Hospital Universitario Parc Taulí); Ines de Mir (Hospital Vall d’Hebron); Gimena Hernandez, Camila Maroni (CAP La Sagrera); Alberto Bercedo (Centro de Salud Los Castros); Jose Antonio Castillo (Hospital Infantil Universitario Miguel Servet); María Teresa Guerra (Centro de Salud Jerez Sur), Olga Cortés (Centro de Salud Canillejas); Eva Tato (Hospital Universitario Araba); Pilar Ortiz, Marta Ortega, Alberto Servan (Centro de Salud Dos de Mayo), María Ángela Carrasco (Consultorio Sevilla la Nueva); Alexandra L. Dima (Institut de Recerca Sant Joan de Déu); Eric van Ganse (University Claude Bernard Lyon); Marijn de Bruin (Radboud UniversityMedical Center).

## Data Availability Statement

Data presented in this study are available upon reasonable request to the corresponding authors.

