## Supplementary Tables for "Impact of Treatment Adherence and Inhalation Technique on asthma outcomes of Pediatric Patients: A Longitudinal Study"

### *Supplementary Material*

#### S1. Multilevel models of asthma symptom control (linear) and exacerbation occurrence (logistic)

| | Asthma symptom control | | Exacerbation occurrence | |
|---|---|---|---|---|
| | Model A | Model B | Model A | Model B |
| | b (SE) | | OR (SE) | |
| **Intercept** | 0.206 (0.221) | 0.084 (0.177) | 0.49 (0.54) | 1.30 (0.60) |
| **Time (years)** | 0.003 (0.003) | 0.001 (0.002) | 0.98 (0.01) * | 0.98 (0.01) ** |
| **ADHERENCE** | | | | |
| **Average adherence** | 0.002 (0.002) | 0.001 (0.002) | 1.00 (0.01) | 0.99 (0.01) |
| **Current fluctuation of adherence** | 0.001 (0.001) | 0.001 (0.001) | 1.00 (0.00) | 1.00 (0.00) |
| **Prior fluctuation of adherence** | -0.003 (0.001) * | -0.001 (0.001) | 0.99 (0.00) | 0.99 (0.00) |
| **INHALATION TECHNIQUE** | | | | |
| **Average IT** | 0.039 (0.038) | 0.011 (0.030) | 1.17 (0.09) | 1.15 (0.10) |
| **Current fluctuation of IT** | 0.014 (0.035) | -0.003 (0.031) | 0.99 (0.11) | 0.96 (0.11) |
| **Prior fluctuation of IT** | -0.036 (0.040) | -0.045 (0.035) | 1.16 (0.12) | 1.15 (0.13) |
| **Treatment** | | | | |
| ICS plus LABA | Ref. | Ref. | Ref. | Ref. |
| ICS | 0.127 (0.088) | 0.091 (0.071) | 0.93 (0.22) | 0.84 (0.23) |
| **Sex** | | | | |
| Male | Ref. | Ref. | Ref. | Ref. |
| Female | 0.173 (0.082) * | 0.133 (0.064) * | 0.97 (0.19) | 0.82 (0.20) |
| **Age** | | | | |
| < 8 years | Ref. | Ref. | Ref. | Ref. |
| 8 – 11 | -0.079 (0.101) | -0.045 (0.080) | 0.43 (0.23) *** | 0.39 (0.24) *** |
| ≥12 | -0.038 (0.115) | -0.003 (0.091) | 0.38 (0.26) *** | 0.31 (0.28) *** |
| **Reliever use** | | | | |
| Almost never | | Ref. | | Ref. |
| Usually | | 0.876 (0.061) *** | | 3.30 (0.21) *** |
| **Exacerbation** | | | | |
| No | | Ref. | | |
| Yes | | 0.327 (0.064) *** | | |
| **Asthma symptom control** | | | | |
| Not well controlled | | | | Ref. |
| Intermediate | | | | 0.33 (0.35) ** |
| Well controlled | | | | 0.37 (0.25) *** |
| ICC (linear); VPC (logistic) | 0.2332 | 0.1498 | 0.0112 | 0.0109 |
| logLikelihood | -1194.0 | -1074.5 | | |
| AIC | 2418.0 | 2183.1 | | |
| BIC | 2489.9 | 2264.5 | | |
| -2 Res Log Pseudo-Likelihood | | | 4149.65 | 4291.46 |
| Generalized Chi-Square | | | 751.82 | 748.04 |
| Generalized Chi-Square / DF | | | 0.85 | 0.85 |

The p-values corresponding to each coefficient or OR provided by the models were marked with asterisks: *(p<0.05); **(p<0.01); ***(p<0.001).

ANOVA p-values for each independent variable were marked with § ($p<0.05$). ICS: Inhaled Corticosteroids; ICS plus LABA: Inhaled Corticosteroids plus Long-Acting Beta-Agonists; ICC: Intraclass Correlation Coefficient; VPC: Variance Partition Coefficients; AIC: Akaike Information Criterion; BIC: Bayesian Information Criterion.

**S2. Multilevel models of Health-Related Quality of Life measured with the EQ-5D and PROMIS-PAIS (linear)**

| | EQ-5D | | PROMIS-PAIS | |
|---|---|---|---|---|
| | Model A | Model B | Model A | Model B |
| | b (SE) | | b (SE) | |
| **Intercept** | 0.964 (0.038) *** | 0.970 (0.040) *** | 11.425 (2.006) *** | 12.441 (2.028) *** |
| **Time (years)** | 0.000 (0.001) | 0.000 (0.001) | -0.026 (0.028) | -0.023 (0.027) |
| **ADHERENCE** | | | | |
| **Average adherence** | 0.000 (0.000) | 0.000 (0.000) | 0.023 (0.020) | 0.023 (0.018) |
| **Current fluctuation of adherence** | 0.000 (0.000) | 0.000 (0.000) | 0.000 (0.009) | -0.004 (0.009) |
| **Prior fluctuation of adherence** | 0.000 (0.000) | 0.000 (0.000) | -0.015 (0.010) | -0.006 (0.010) |
| **INHALATION TECHNIQUE** | | | | |
| **Average IT** | 0.005 (0.006) | 0.006 (0.006) | -0.528 (0.356) | -0.558 (0.334) |
| **Current fluctuation of IT** | -0.013 (0.006) * | -0.012 (0.006) * | -0.323 (0.316) | -0.363 (0.312) |
| **Prior fluctuation of IT** | 0.017 (0.007) * | 0.017 (0.007) * | -0.054 (0.350) | 0.051 (0.346) |
| **Treatment** | | | | |
| ICS plus LABA | Ref. | Ref. | Ref. | Ref. |
| ICS | -0.001 (0.016) | -0.001 (0.016) | -0.133 (0.863) | -0.033 (0.814) |
| **Sex** | | | | |
| Male | Ref. | Ref. | Ref. | Ref. |
| Female | -0.032 (0.014) * | -0.033 (0.014) * | 2.384 (0.779) ** | 2.512 (0.729) *** |
| **Age** | | | | |
| < 8 years | Ref. | Ref. | Ref. | Ref. |
| 8 - 11 | 0.001 (0.017) | -0.004 (0.017) | -0.400 (0.931) | -0.140 (0.879) |
| ≥ 12 | 0.002 (0.020) | -0.002 (0.020) | 2.462 (1.111) * | 2.727 (1.051) * |
| **Asthma symptom control** | | | | |
| Not well controlled | | Ref. | | Ref. |
| Intermediate | | -0.006 (0.018) | | 0.181 (0.995) |
| Well controlled | | 0.009 (0.014) | | -2.250 (0.800) ** |
| **Reliever use** | | | | |
| No | | Ref. | | Ref. |
| Yes | | -0.017 (0.011) | | 1.386 (0.621) * |
| **Exacerbation** | | | | |
| No | | Ref. | | Ref. |
| Yes | | -0.018 (0.011) | | -0.154 (0.570) |
| ICC (linear) | 0.6223 | 0.6192 | 0.5512 | 0.5130 |
| logLikelihood | 350.6 | 341.8 | -1518.3 | -1518.3 |
| AIC | -667.2 | -641.5 | 3104.2 | 3078.6 |
| BIC | -596.6 | -554.5 | 3175.4 | 3166.3 |

The p-values corresponding to each coefficient provided by the models were marked with asterisks: *(p<0.05); **(p<0.01); ***(p<0.001).
ANOVA p-values for each independent variable were marked with § (p<0.05).
ICS: Inhaled Corticosteroids; ICS plus LABA: Inhaled Corticosteroids plus Long-Acting Beta-Agonists; ICC: Intraclass Correlation Coefficient; VPC: Variance Partition Coefficients; AIC: Akaike Information Criterion; BIC: Bayesian Information Criterion.
